# Loss of Peripheral Retinal Vessels in Retinitis Pigmentosa

**DOI:** 10.1101/2024.10.18.24315764

**Authors:** Hossein Ameri, Alexander T. Hong, Jason Chwa

## Abstract

**Objective:** Retinitis pigmentosa (RP) is the most common inherited retinal disease and a major cause of irreversible vision loss. The purpose of this study was to assess peripheral retinal vessels in RP.

**Design:** Cross-sectional study

**Subjects:** Patients with RP and age-matched controls.

**Methods:** Using ultra-wide field fundus images, the retina was divided into three zones: posterior, mid periphery, and far periphery. To evaluate vascularity of the retina, the vessels were counted at the border of posterior and mid peripheral zones (Z1/2) and the border of mid peripheral and far peripheral zones (Z2/3).

**Main outcome measures:** Vessel counts at Z1/2 and Z2/3

**Results:** 181 eyes of 107 RP patients and 130 eyes of 84 controls were included. In the RP group, the median vessel counts at Z1/2 and Z2/3 were 8 and 3, respectively. These were strikingly lower than the control group with the median vessels of 42 and 43.5 at Z1/2 (p<0.001) and Z2/3 (p<0.001), respectively. 22% of RP eyes were entirely avascular in the far periphery and 7% were avascular in the mid periphery and far periphery. Only 5% of RP eyes had more than 25 vessels at Z2/3. There were significantly fewer vessels in the temporal retina at both Z1/2 (p=0.01) and Z2/3 (p<0.001) in RP eyes. Furthermore, eyes with visual acuity of 20/200 or worse had significantly fewer vessels at Z1/2 (p<0.001) and Z2/3 (p<0.001). There were no significant differences in the number of vessels at Z1/2 and 2/3 between the right and left eyes of RP patients with both eyes included in the study.

**Conclusions:** This study provides compelling evidence of substantial symmetrical peripheral retinal vascular loss in RP. This finding may aid in clinical diagnosis of the disease and have significant therapeutic implications.

---

Retinitis pigmentosa (RP) is the most common cause of hereditary vision loss with a worldwide prevalence of 1/3000 to 1/5000 ^1,2^. It generally manifests with night blindness and subsequently progresses to peripheral vision loss, ultimately leading to loss of central vision. The earliest accounts of fundus observations and retinal histopathology in RP date back to the mid-19th century ^3,4^. However, familial night blindness was described as early as the mid-18th century, and a combination of impaired vision and retinal pigmentation was reported in the early 19th century ^4^. RP is primarily a disease of rod photoreceptors and cones are often secondarily affected. In a typical RP, fundus exam shows bone spicule pigmentation, vascular attenuation, and waxy pallor of the disc. However, some patients may show only one or two of these signs at a given time, and in early stages of the disease, fundus exam may appear normal. In recent years, with the advent of optical coherence tomography angiography (OCTA), retinal and choroidal vasculature have been studied in more detail. However, studies of the retinal vasculature have been limited to the posterior pole only. We have clinically observed a lack of vascularity of the peripheral retina in RP. The goal of this study was to examine this clinical observation in detail.

## Methods

This is a retrospective cross-sectional study conducted in an academic tertiary referral center. The study design adhered to the Declaration of Helsinki and was approved by the Institutional Review Board of the University of Southern California.

### Patient Selection

The study population consisted of RP patients and age-matched control patients seen at the USC Roski Eye Institute between April 2016 and April 2024. For the RP group, only patients with a clear diagnosis of syndromic or non-syndromic RP were included (Fig. 1). Patients with any history of retinal vascular disease, inflammation, retinal detachment, or any pathology that could potentially alter the structure of the retinal periphery or vessels were excluded.

**Figure 1.**
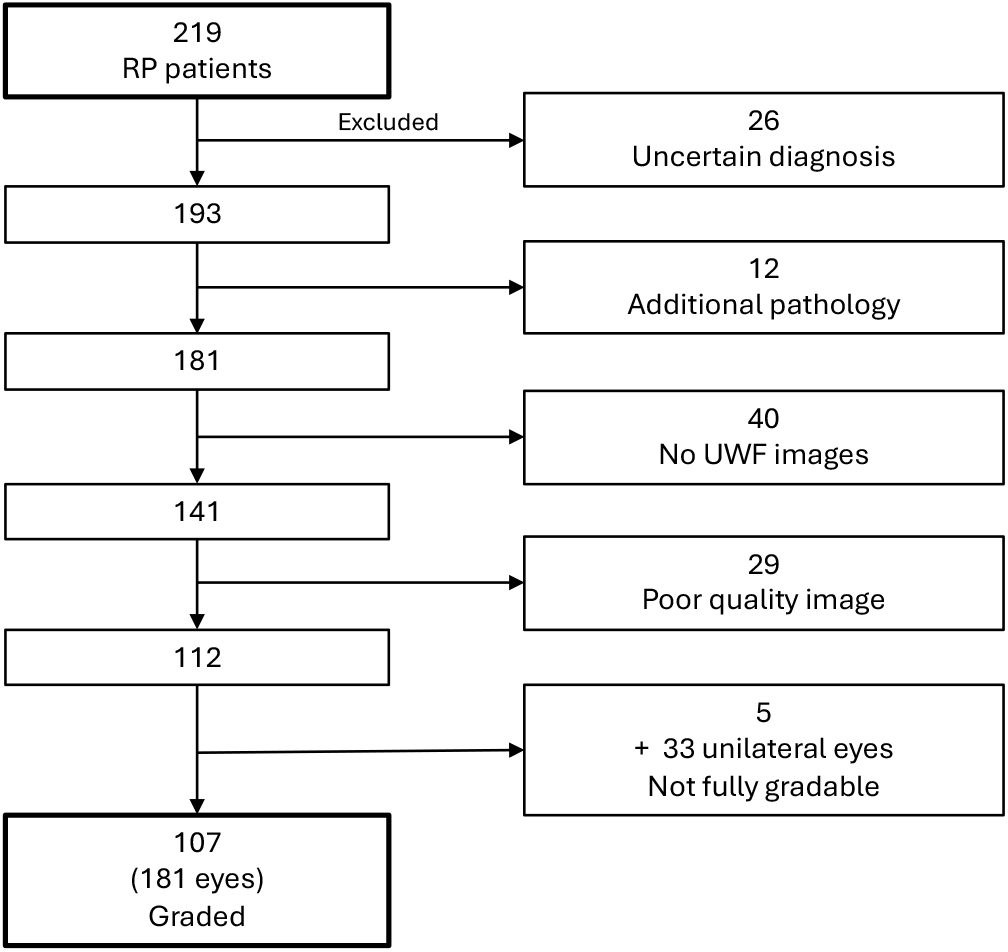
Flowchart diagram showing the process of selecting images of patients with RP.

### Vessel Count on Fundus Images

Ultra-wide field (UWF) images obtained by a fundus camera (Optos California FA/ICG, Marlborough, MA) were used for the analysis of retinal vessels. Only eyes with good quality UWF images and visible retinal periphery were included, and the best image with the most area exposure and clarity was selected (Fig. 1). For fluorescein angiogram (FA), images between one and three minutes with the highest visibility of vessels were selected. The retina was divided into four quadrants by drawing a vertical and a horizontal line crossing the center of the optic disc (Fig. 2). The retina was also divided into three zones, zone 1 (posterior), zone 2 (mid periphery), and zone 3 (far periphery) by drawing 2 concentric circles, centered on the horizontal line at the fovea level. The radius of the small circle, which marked the border between zone 1 and zone 2 (Z1/2) was twice the distance between the fovea and the center of the disc. The radius of the large circle, which marked the border between zone 2 and zone 3 (Z2/3) was three times the distance between the fovea and the center of the disc. Vessels crossing Z1/2 and Z2/3 were counted by two independent observers and used as a vascularity index. In eyes with a discrepancy between the two counts, a third observer counted the vessels and determined the final count.

**Figure 2.**
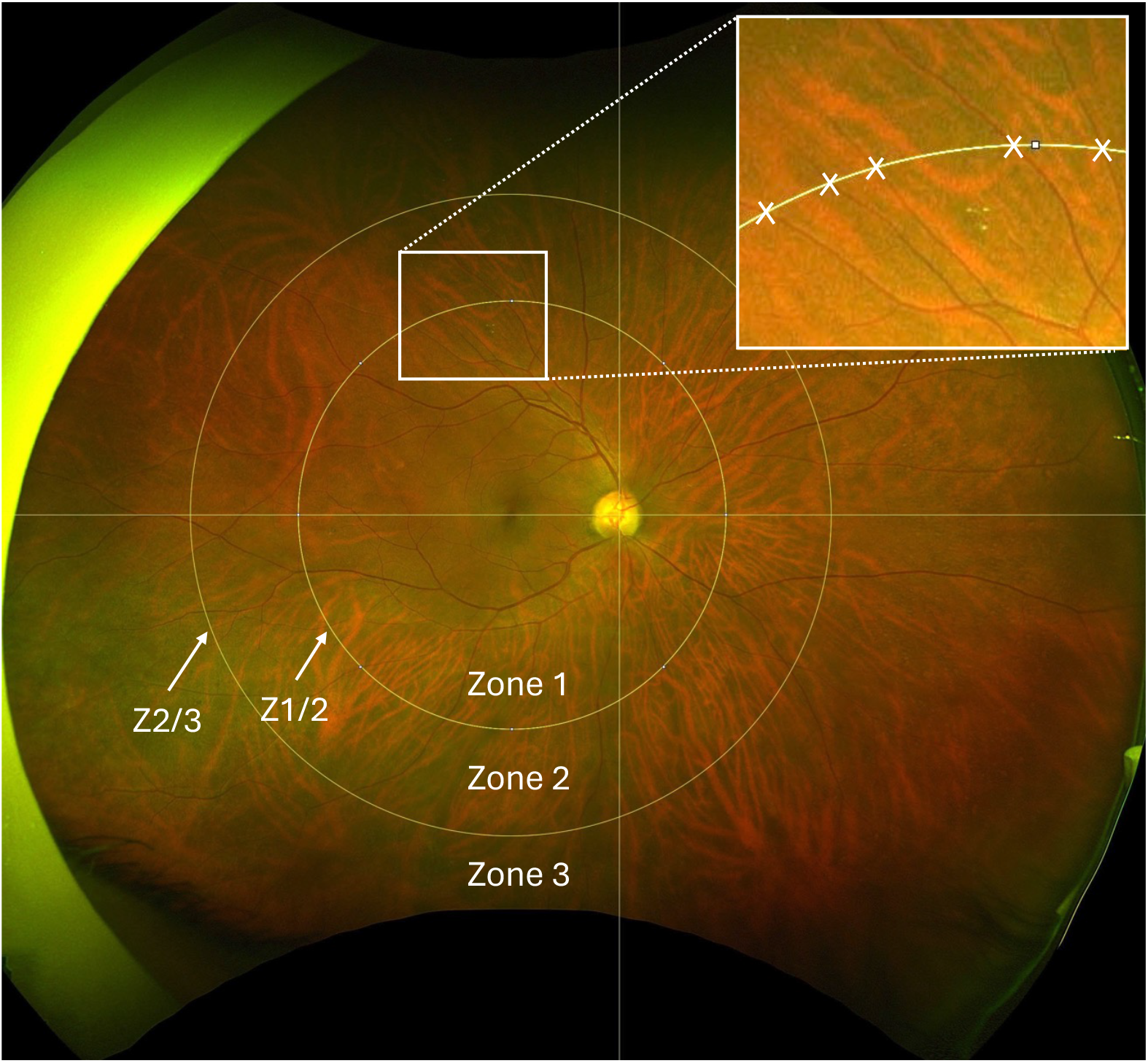
Representative ultra-wide field fundus photo of a control eye showing Z1/2 and Z2/3 rings. The inset on the top right is a magnified portion of the superotemporal Z1/2 with crossing vessels marked by X.

### Statistical Analysis

Descriptive statistics were used to summarize the baseline demographic characteristics. Student’s t-test was used to explore differences between continuous variables and the Chi-squared test was used for categorical variables. Normality was assessed using the Shapiro-Wilk test. Data were analyzed by sub-cohorts: 1) Zone-based analysis: To investigate the differences in mean vessel count between the RP and Control groups, regression models were fitted separately for Z1/2 and Z2/3. Models were fitted for outcomes of mean vessel count per eye, mean vessel count per quadrant, and mean vessel count per hemisphere. 2) Factor-based analysis: To compare the adjusted incidence rate ratios (IRR), models were fit separately for each control and RP group. 3) Visual Acuity (VA) Analysis: Snellen VA was converted to Logarithmic Minimum Angle of Resolution (logMAR), and initially used as a continuous variable. To evaluate the linearity assumption between VA and total vessel counts, beta coefficients were extracted and plotted from a model treating VA as a categorical variable and divided into 11 different groups; this is the highest categorization we could divide to remain model convergent. Non-linearity was identified from visual inspection of beta estimate plots (Supplementary Figure 1). We then explored multiple data transformations, which include quartiles, two or three categorizations based on visual inspection, quadratic and cubic polynomials, and splines with two and three segments. The final model was selected based on the clinically relevant cut-off and Quasi-Information Criterion (QIC). The final model selected was three categorizations with the logMAR VA cut-offs at 0.48 (Snellen equivalent of 20/60) and 1.0 (Snellen equivalent of 20/200). Another similar QIC model was three categorizations with the logMAR VA cut-offs at 0.7 (Snellen equivalent of 20/100) and 1.0. Since 0.48 is more clinically relevant, we used the 0.48 cut-off model as our main result. We also present the 0.7 cut-off model as a sensitivity analysis in the appendix.

To account for bilateral eye data, generalized estimating equations (GEE) were used. Poisson regression was used for comparison between Z1/2 and Z2/3, and negative binomial regression was used for all other outcomes to account for variance inflation. The GEEs were fit using an exchangeable correlation structure to account for within-subject correlations. The correlation structure was chosen by the QIC. Results are reported as IRR with 95% confidence interval (CI) for all outcomes. For the analysis of FA images, paired differences between FA and color counts were calculated for each zone. For each participant, differences in counts were computed, and the mean and standard deviation of these differences were calculated. Paired Wilcoxon signed-rank tests were performed to assess the significance of differences between FA and color counts. Statistical significance was set at p<0.05, 2-sided. All statistical analyses were conducted using R software (version 4.3.3, R Foundation for Statistical Computing, Vienna, Austria).

## Results

The study included 181 eyes of 107 patients with RP and 130 eyes of 84 individuals with no major retinal pathology as controls (Table 1). Table 2 shows the mean number of vessel counts at Z1/2 and Z2/3 in the RP and control eyes. In the RP group, the median number of vessels at Z1/2 and Z2/3 were 8 and 3, respectively. These were markedly lower than the control group with the median vessels of 42 and 43.5 at Z1/2 and Z2/3, respectively (Fig. 3, Fig. 4).

**Table 1.** Baseline demographic characteristics of the study participants. Age is presented as average ± standard deviation.

| Variable | Control<br>(n=84) | Retinitis Pigmentosa<br>(n=107) | P Value | Total<br>(n=191) |
| --- | --- | --- | --- | --- |
| Age, years | 48.3 ± 17.4<br>(range 14 – 86) | 47 ± 18.3<br>(range 19 – 86) | 0.62 | 47.6 ± 17.9 |
| Sex |  |  | 0.07 |  |
| Female | 48 (57%) | 46 (43%) |  | 94 (49%) |
| Male | 36 (43%) | 61 (57%) |  | 97 (51%) |

**Table 2.** Average vessel counts (± standard deviation) at Z1/2 and Z2/3 by quadrants and hemispheres in the control and retinitis pigmentosa groups. IN: inferonasal, IT: inferotemporal, SN: superonasal, ST: superotemporal

|  | Control<br>(n=130) |  | Retinitis Pigmentosa<br>(n=181) |  |
| --- | --- | --- | --- | --- |
|  | Z1/2 | Z2/3 | Z1/2 | Z2/3 |
| Total | 42.8 ± 6.7 | 43.9 ± 7.7 | 10.4 ± 9.4 | 6.5 ± 8.7 |
| ST | 13.3 ± 2.7 | 12 ± 3 | 2.8 ± 3.4 | 1.4 ± 3.1 |
| IT | 12.9 ± 3.7 | 13.3 ± 3.6 | 2 ± 3.1 | 1 ± 2.6 |
| SN | 8.9 ± 1.9 | 9.7 ± 2.1 | 3.2 ± 2.4 | 2.4 ± 2.5 |
| IN | 7.8 ± 1.8 | 8.9 ± 2.3 | 2.5 ± 1.9 | 1.8 ± 2.1 |
| Temporal | 26.1 ± 4.8 | 25.3 ± 5.4 | 4.8 ± 6.1 | 2.3 ± 5.4 |
| Nasal | 16.7 ± 3.1 | 18.7 ± 3.5 | 5.7 ± 4 | 4.2 ± 4.4 |
| Superior | 22.1 ± 3.8 | 21.7 ± 4.3 | 6 ± 5.4 | 3.8 ± 5 |
| Inferior | 20.7 ± 4.3 | 22.2 ± 5 | 4.5 ± 4.5 | 2.7 ± 4.2 |

**Figure 3.**
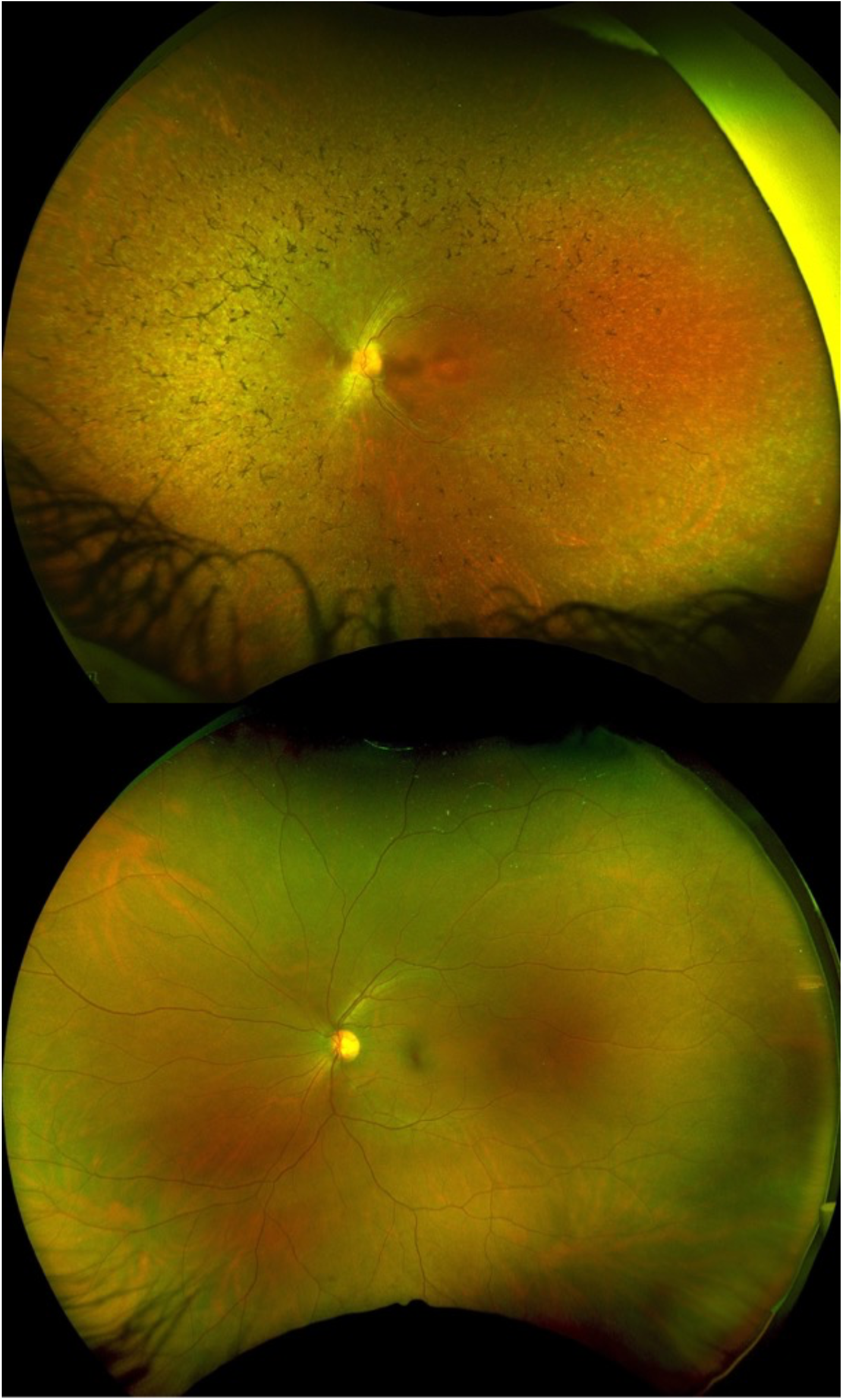
Representative ultra-wide field fundus photo of an RP eye (top) and a control eye (bottom). Note extensive vessel loss in the periphery of the RP eye. The vessel counts in the RP eye were 5 and 3 at Z1/2 and Z2/3, respectively; these numbers in the control eye were 41 and 50, respectively.

**Figure 4.**
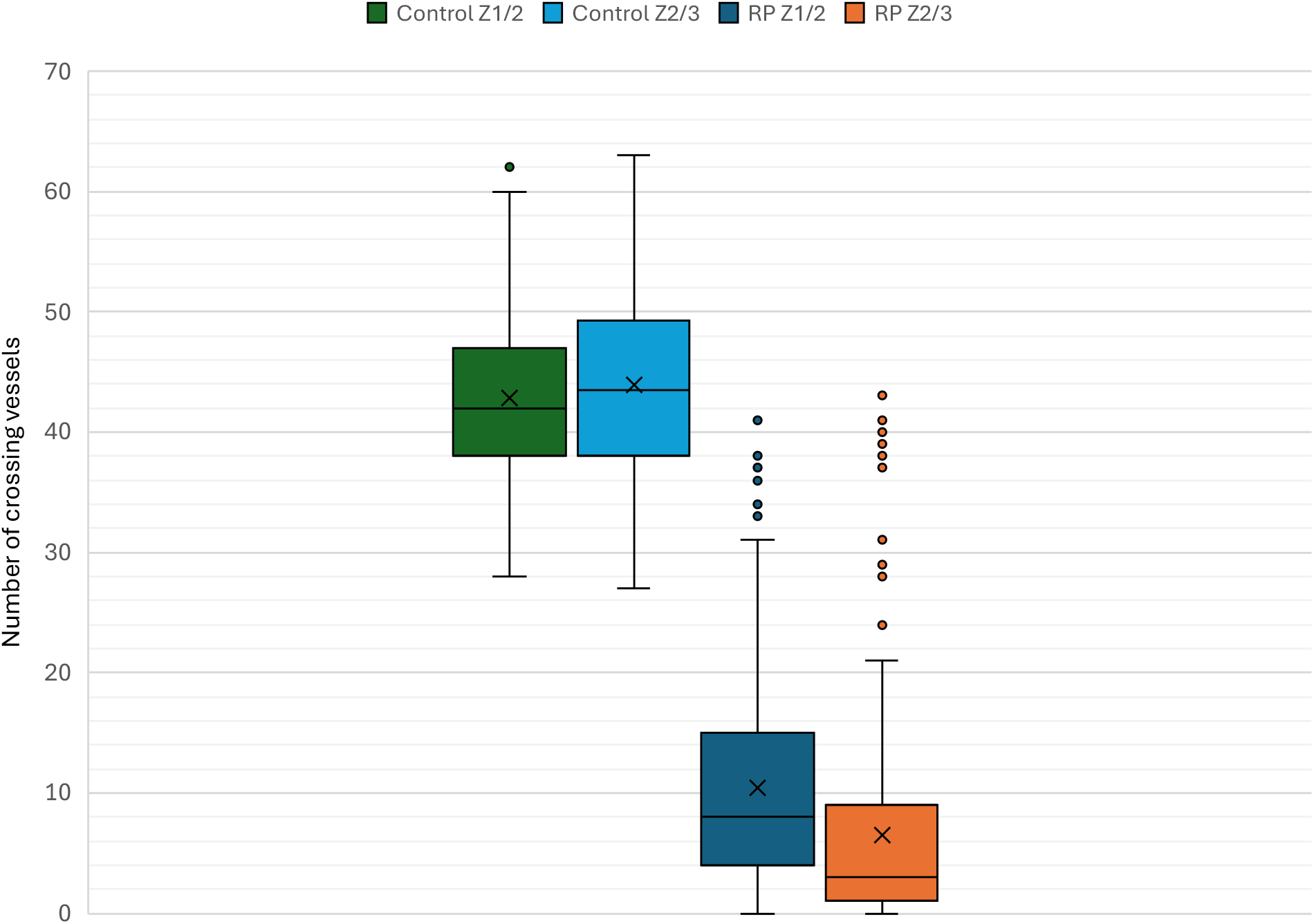
Box and whisker plot of control and retinitis pigmentosa (RP) eyes for Z1/2 and Z2/3. The horizontal line in each box denotes the median and X represents the mean.

To compare the vessel count in the RP eyes with the control eyes, GEE models controlling for age and sex examined differences between vessel count at Z1/2 and Z2/3 (Table 3). Vessel count was significantly lower in both Z1/2 (p < 0.001) and Z2/3 (p < 0.001) in RP patients. There was an association between the age and the vessel count; there were significantly fewer vessels in the age quartile 64-86 at both Z1/2 (p<0.001) and Z2/3 (p=0.001). Sex did not significantly influence overall vessel count (Z1/2: p=0.37; Z2/3: p=0.65).

**Table 3.** Comparison of adjusted incidence rate ratio (IRR) for the control and retinitis pigmentosa (RP) groups at Z1/2 and Z2/3. CI: confidence interval, REF: reference

|  | Z1/2 |  |  | Z2/3 |  |  |
| --- | --- | --- | --- | --- | --- | --- |
| Variable | IRR | 95% CI | P Value | IRR | 95% CI | P Value |
| Factor *<br>disease |  |  |  |  |  |  |
| Control | REF |  |  | REF |  |  |
| RP | 0.24 | 0.20, 0.28 | <0.001 | 0.14 | 0.11, 0.18 | <0.001 |
| Age |  |  |  |  |  |  |
| 14-33 | REF |  |  | REF |  |  |
| 34-47 | 0.99 | 0.77, 1.26 | 0.93 | 1.21 | 0.86, 1.71 | 0.27 |
| 48-63 | 0.77 | 0.61, 0.97 | 0.02 | 0.86 | 0.62, 1.18 | 0.34 |
| 64-86 | 0.66 | 0.54, 0.82 | <0.001 | 0.63 | 0.48, 0.82 | 0.001 |
| Sex |  |  |  |  |  |  |
| Female | REF |  |  | REF |  |  |
| Male | 1.09 | 0.91, 1.30 | 0.37 | 1.06 | 0.82, 1.37 | 0.65 |

In the RP group, in 13 eyes (7%) there were no vessels at Z1/2 and the entire peripheral retina beyond Z1/2 was avascular (Fig. 5). The maximum number of vessels at Z1/2 was 41. In 39 eyes (22%) there were no vessels at Z2/3, and the peripheral retina beyond Z2/3 was avascular. In 26 eyes (14%) there were one or more vessels at Z1/2 but zero vessels at Z2/3. The maximum number of vessels at Z2/3 was 43. In 119 eyes (66%) there were 10 or fewer vessels at Z1/2 and only in 16 eyes (9%) were there more than 25 vessels. In 149 eyes (82%) there were 10 or fewer vessels at Z2/3 and only in 9 eyes (5%) were there more than 25 vessels.

**Figure 5.**
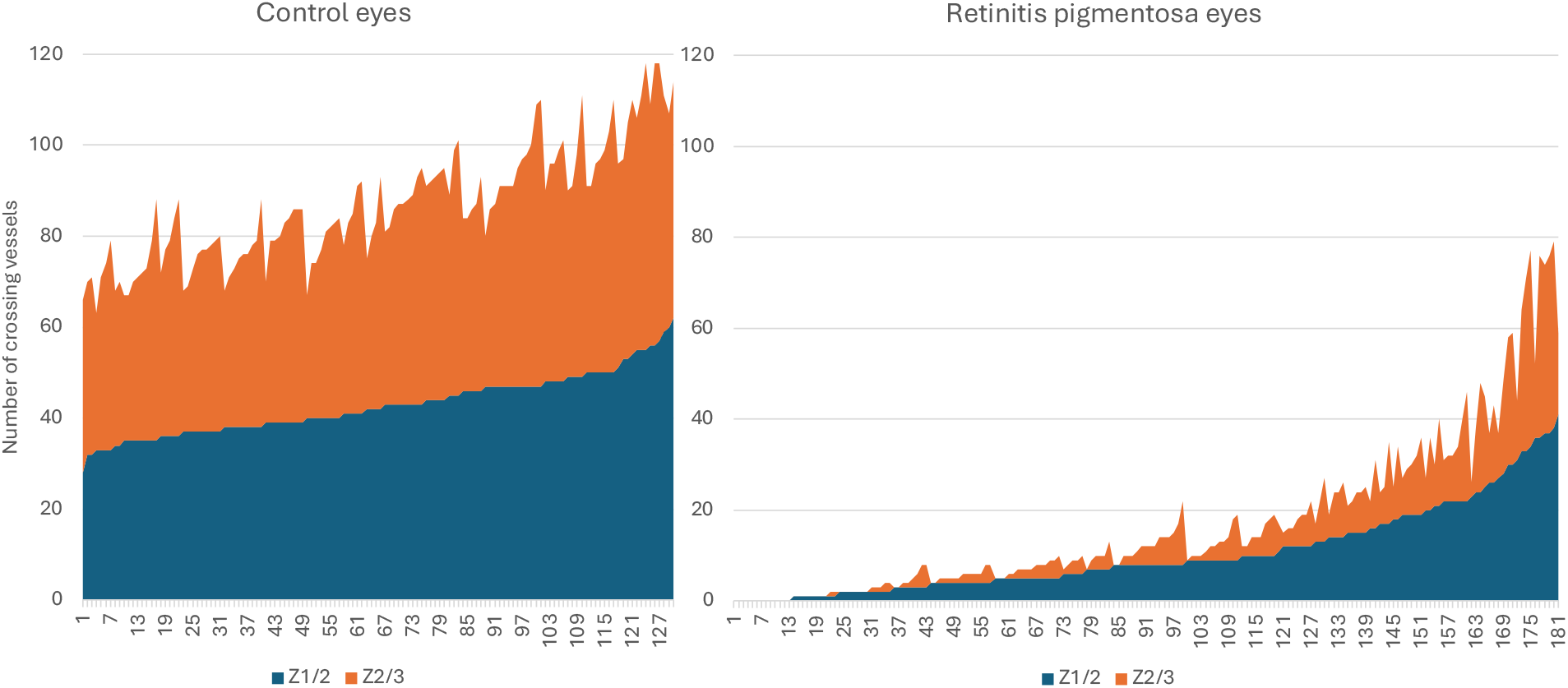
Bar graph showing all vessel counts in the control (left) and RP (right) eyes. Each number on the X axis represents one eye. For each eye, the Z2/3 count (orange) has been stacked on the Z2/3 count (blue). Note the striking difference between the control and RP eyes.

There were significantly fewer vessels at Z2/3 compared to Z1/2 (p<0.001) (Table 4). The mean and median differences were 3.91 (95% CI 2.03, 5.78), and 3, respectively. In 24 eyes the number of vessels was the same at Z1/2 and Z2/3. In 144 eyes there were more vessels at Z1/2 than Z2/3, ranging from 1 to 23 vessels, and in 13 eyes there were more vessels at Z2/3 than Z1/2, ranging from 1 to 9 vessels.

**Table 4.**
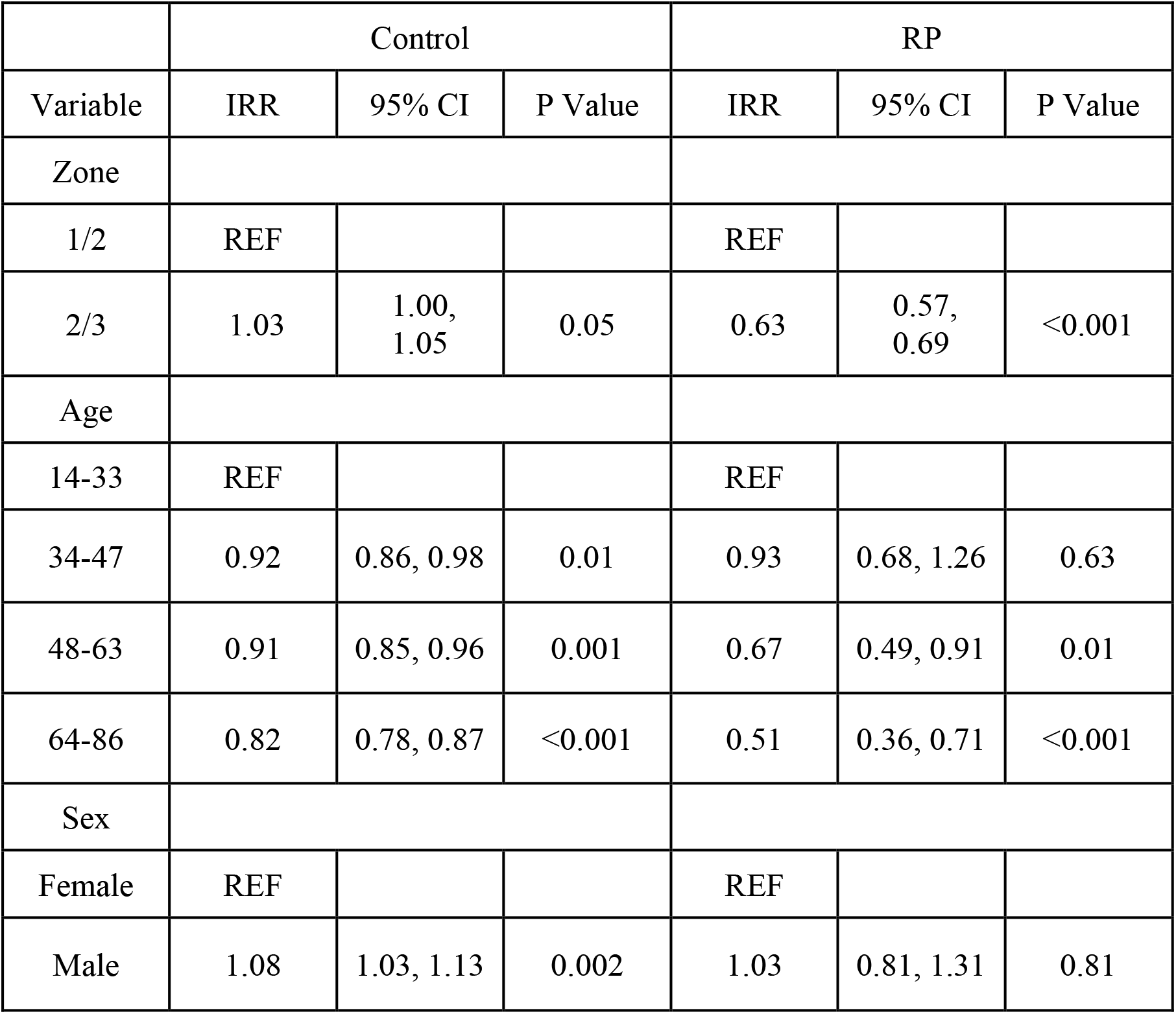
Comparison of adjusted incidence rate ratio (IRR) for Z1/2 and Z2/3 in the control and retinitis pigmentosa (RP) groups. CI: confidence interval, REF: reference

|  | Control |  |  | RP |  |  |
| --- | --- | --- | --- | --- | --- | --- |
| Variable | IRR | 95% CI | P Value | IRR | 95% CI | P Value |
| Zone |  |  |  |  |  |  |
| 1/2 | REF |  |  | REF |  |  |
| 2/3 | 1.03 | 1.00, 1.05 | 0.05 | 0.63 | 0.57, 0.69 | <0.001 |
| Age |  |  |  |  |  |  |
| 14-33 | REF |  |  | REF |  |  |
| 34-47 | 0.92 | 0.86, 0.98 | 0.01 | 0.93 | 0.68, 1.26 | 0.63 |
| 48-63 | 0.91 | 0.85, 0.96 | 0.001 | 0.67 | 0.49, 0.91 | 0.01 |
| 64-86 | 0.82 | 0.78, 0.87 | <0.001 | 0.51 | 0.36, 0.71 | <0.001 |
| Sex |  |  |  |  |  |  |
| Female | REF |  |  | REF |  |  |
| Male | 1.08 | 1.03, 1.13 | 0.002 | 1.03 | 0.81, 1.31 | 0.81 |

In the control group, the median number of vessels at Z1/2 and Z2/3 were 42 and 43.5, respectively. The difference between the Z1/2 and Z2/3 vessels was not statistically significant (p=0.05).

Comparing the quadrants (Table 5, Supplementary Table 1), in RP patients, there were significantly more vessels in the superonasal quadrant at both Z1/2 (p<0.001) and Z2/3 (p<0.001), and significantly fewer vessels in the inferotemporal quadrant at Z2/3 (Z1/2: p=0.05; Z2/3: p=0.001). In the control group, there were significantly more vessels in the superotemporal quadrant at Z1/2 (p<0.001) and the inferotemporal quadrant at Z2/3 (p<0.001). There were significantly fewer vessels in the inferonasal quadrant at both Z1/2 (p<0.001) and Z2/3 (p=0.002).

**Table 5.**
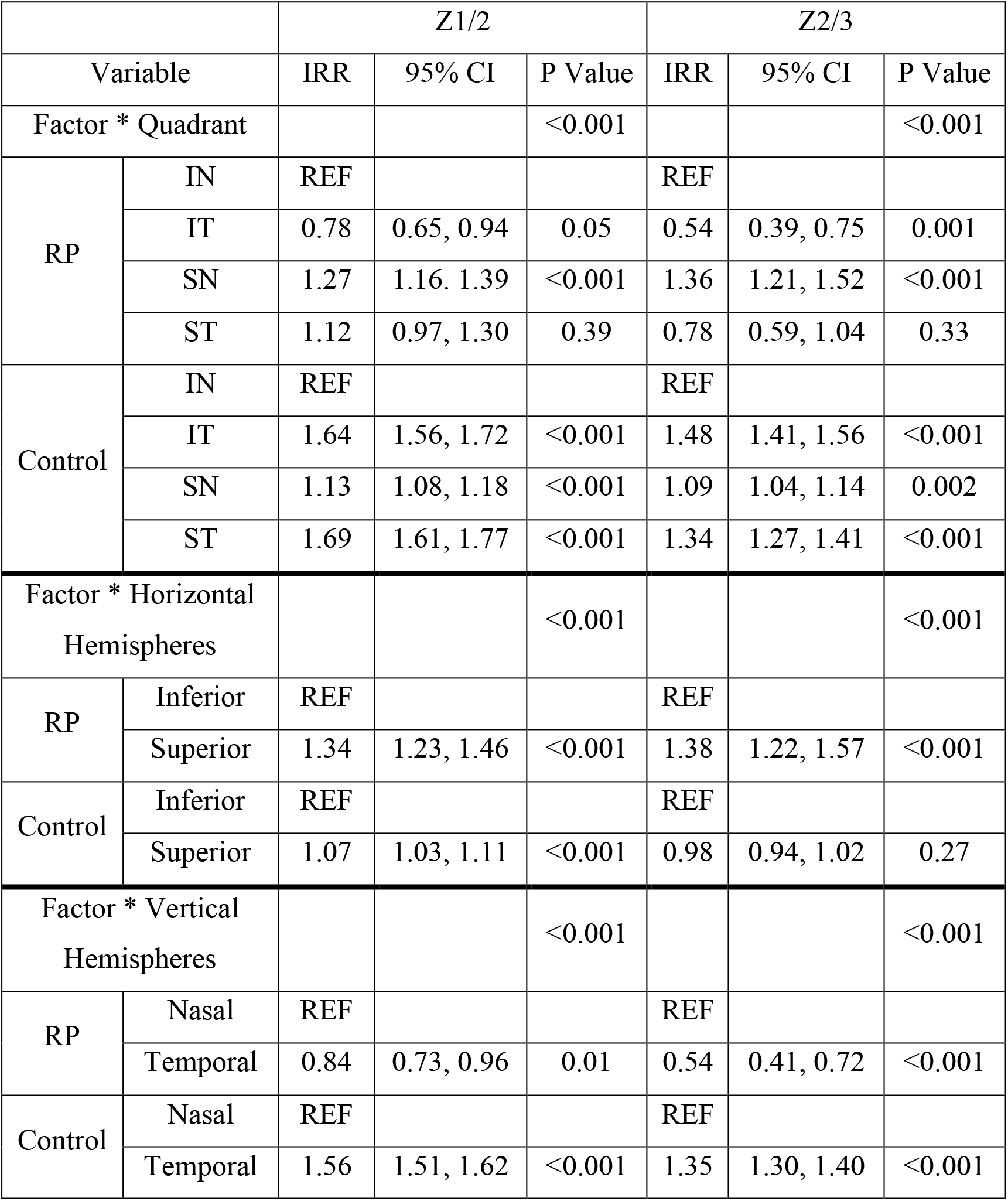
Comparison of adjusted incidence rate ratio (IRR) for quadrants and horizontal and vertical hemispheres at Z1/2 and Z2/3 in the control and retinitis pigmentosa (RP) groups. For more detailed analysis, refer to supplementary tables 1, 2, and 3. CI: confidence interval, REF: reference, IN: inferonasal, IT: inferotemporal, SN: superonasal, ST: superotemporal

Comparing the superior and inferior retinal hemispheres, (Table 5, Supplementary Table 2), while there were significantly more vessels in the superior retina at both Z1/2 (p<0.001) and Z2/3 (p<0.001) in RP patients, and at Z1/2 in the control group (p<0.001), there was no difference between the superior and inferior retina at Z2/3 in the control group (p=0.27). When comparing the temporal and nasal retinal hemispheres (Table 5, Supplementary Table 3), there were significantly fewer vessels in the temporal retina at both Z1/2 (p=0.01) and Z2/3 (p<0.001) in RP patients. In contrast, there were significantly more vessels in the temporal retina at both Z1/2 (p<0.001) and Z2/3 (p<0.001) in control eyes.

To assess how symmetric the vessel loss in RP is, the right and left eyes were compared in 74 RP patients who had images for both eyes (Supplementary Table 4). The mean vessels at Z1/2 were 10.65 for the right and 10.15 for the left eyes. There was no significant difference between the two eyes (p=0.39). The mean vessels at Z2/3 were 6.30 for the right and 6.57 for the left eyes. There were no statistically significant differences between the two eyes (p=0.56). In 46 control patients with images of both eyes included in the study (Supplementary Table 5), the crossings were slightly higher in the right eye for Z1/2 (44.4 ± 7.0 vs 42.5 ± 6.6, p=0.03) but the same for Z2/3 (44.9 ± 7.2 vs 44.9 ± 7.7, p=1.00).

To determine whether VA correlates with vessel counts (Table 6, Supplementary Table 6), VA was collected for all but two eyes of one patient whose VA on the day of imaging was not available. There were no significant differences in baseline logMAR VA between right and left eyes (right: 1.14 ± 1.08; left:1.27 ± 1.07, p=0.43). Given non-linearity of the relationship between logMAR VA and vessel counts (Supplementary Fig. 1), logMAR VAs were divided into three categories: 0-0.48, 0.49-0.99, and 1-3. After adjusting for age quartiles and sex, the logMAR VA 1.0-3.0 group (Snellen equivalent of worse than 20/200) showed significantly fewer vessel counts at Z1/2 (p < 0.001) and Z2/3 (p < 0.001) compared to the logMAR VA 0-0.48 group (Snellen equivalent of 20/20 to 20/60).

**Table 6.**
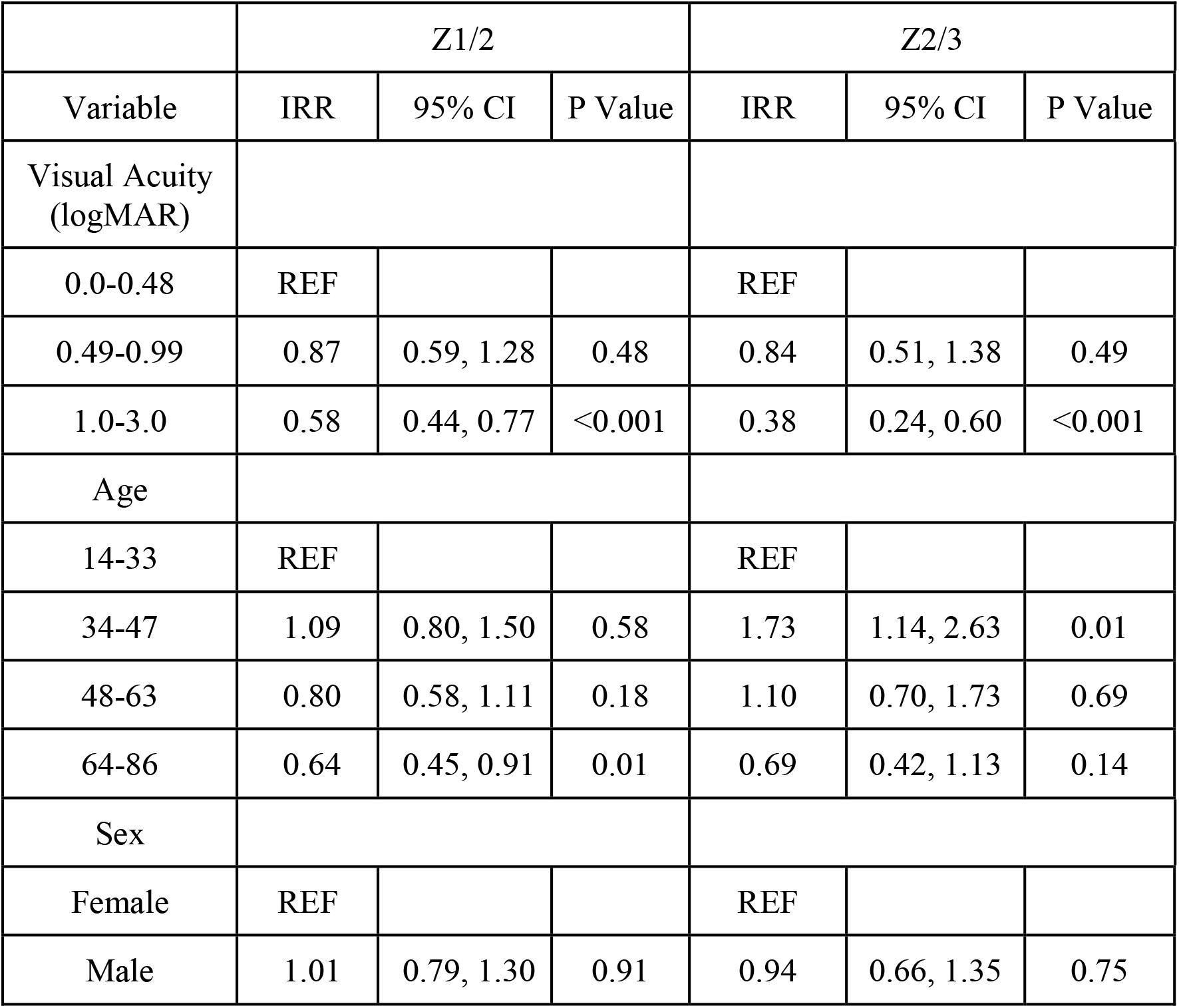
Comparison of adjusted incidence rate ratio (IRR) of logMAR visual acuity in retinitis pigmentosa patients. CI: confidence interval, REF: reference

|  | Z1/2 |  |  | Z2/3 |  |  |
| --- | --- | --- | --- | --- | --- | --- |
| Variable | IRR | 95% CI | P Value | IRR | 95% CI | P Value |
| Visual Acuity (logMAR) |  |  |  |  |  |  |
| 0.0-0.48 | REF |  |  | REF |  |  |
| 0.49-0.99 | 0.87 | 0.59, 1.28 | 0.48 | 0.84 | 0.51, 1.38 | 0.49 |
| 1.0-3.0 | 0.58 | 0.44, 0.77 | <0.001 | 0.38 | 0.24, 0.60 | <0.001 |
| Age |  |  |  |  |  |  |
| 14-33 | REF |  |  | REF |  |  |
| 34-47 | 1.09 | 0.80, 1.50 | 0.58 | 1.73 | 1.14, 2.63 | 0.01 |
| 48-63 | 0.80 | 0.58, 1.11 | 0.18 | 1.10 | 0.70, 1.73 | 0.69 |
| 64-86 | 0.64 | 0.45, 0.91 | 0.01 | 0.69 | 0.42, 1.13 | 0.14 |
| Sex |  |  |  |  |  |  |
| Female | REF |  |  | REF |  |  |
| Male | 1.01 | 0.79, 1.30 | 0.91 | 0.94 | 0.66, 1.35 | 0.75 |

To assess the rate of visibility of vessels on color fundus photos, the color images were compared with the FA images in 21 eyes of 13 RP patients. In this group, significantly more vessels were visible on FA images at Z1/2 (22.2 ± 16.1) compared to the color images (15.6 ± 7.8) (p=0.02). There were also more visible vessels on FA images at Z2/3 (13.7 ± 16) compared to the color images (9.5 ± 6.9) but this was not statistically significant (p=0.05).

## Discussion

This study provides a detailed analysis of peripheral retinal vessels in RP patients using UWF fundus imaging. We devised a method to quantify vessels in the peripheral retina and used vessel crossings at Z1/2 and Z2/3 as a vascularity index. The findings underscore a significant reduction in the number of vessels in the peripheral retina. Analysis of the eyes with gradable imaging for both eyes indicated that this loss is symmetrical. Comparing the vascularity index of the retinal periphery between the RP and control cohorts clearly demonstrates a substantial decline in peripheral retinal perfusion in RP. Furthermore, examination of FA images from a small cohort of patients, showed that the low vascularity index in RP is not because of a lack of visibility of perfusing vessels but rather reflects a real phenomenon. Remarkably in 22% of RP patients, the retina was entirely avascular in the far periphery, and in 82% there were 10 or fewer vessel crossings at Z2/3 compared to the control cohort which had at least 27 vessel crossings in all eyes. The vascular changes in RP were symmetrical. In addition to the vascularity index, there were other differences between the RP and control cohorts. While there was no significant difference between Z1/2 and Z2/3 vessel crossings in the control cohort (average 42.8 and 43.9, respectively), the difference between Z1/2 and Z2/3 vessel crossings in the RP patients (average 10.4 and 6.5, respectively) was highly statistically significant. This difference suggests a gradual loss of retinal vessels from the periphery toward the center. Another interesting difference between the RP and control cohorts was the difference between the nasal and temporal vessel crossings. While the vessel crossings were significantly higher in the temporal side in the control cohort for both Z1/2 and Z2/3, the vessel crossings in the RP cohort showed a lack of difference for Z1/2 and a reverse phenomenon for Z2/3 with the nasal retina showing significantly higher vessel crossings. This is perhaps another indication that the vascular loss progresses from the periphery to the center, and since our circles of Z1/2 and Z2/3 are not centered at the optic disc, they naturally capture a much longer length of temporal vessels compared to the nasal ones (Fig 2).

Our study also demonstrated that patients with VA of worse than 20/200 had a significantly lower number of blood vessels in the retinal periphery. Given that a VA of worse than 20/200 is often seen in the advanced stage of RP, this finding is suggestive of a potential correlation between the peripheral retinal vessels and RP severity.

To the best of our knowledge, this is the first study of the peripheral retinal vasculature in RP patients. However, retinal vessel diameter in the posterior pole has been studied in RP ^5,6^, and it has been shown to be narrower than in healthy individuals ^6^. Moreover, vascular attenuation has been shown to correlate with the severity of visual field loss ^5,6^ and electroretinogram abnormality ^6^. In addition, retinal blood flow velocity has been reported to be significantly lower in both arteries and veins in RP patients compared to healthy subjects ^7,8^. Retinal capillaries have also been extensively studied in the posterior pole of the retina in recent years using optical coherence tomography angiography (OCTA). RP patients have been reported to have decreased vascular density in the superficial and deep capillary plexus and choriocapillaris in the macular region ^9^ as well as in the radial peripapillary capillary network ^10^. Correlations of these findings with visual function and retinal structural changes have also been reported ^9–11^. Our study demonstrated a correlation between the VA and peripheral retinal vessels; future studies could explore potential correlations between the peripheral retinal vessels and other visual functions in RP patients.

Human histopathological studies in RP patients have shown relative preservation of the inner nuclear layer and, to a lesser extent, the ganglion cell layer at the posterior pole ^12^, with a gradual loss of both layers from the center to the periphery in the extramacular region as far as 10 mm from the foveal center ^13^, i.e. Z1/2. Further studies are needed to determine the extent of inner retinal preservation in the mid-periphery and far peripheral retina in RP patients with peripheral retinal vascular loss.

Histological study of the retinal vessels in humans has been limited to the areas of bone spicule formation, which showed very thin endothelial cells with scattered fenestrations, resembling the choriocapillaris ^14^. Retinal vessels have been studied in the mouse and rat models of retinal degeneration, and all have demonstrated overall regression of retinal vessels over time ^15–18^. Although the density of capillaries and small vessels has been reported to decrease ^16–18^, the report on large retinal vessels has been mixed, with no major change ^18^, or a milder reduction compared to small vessels ^16,18^. None of these animal studies have shown avascular peripheral retinas resembling the RP patients in our study. Larger animal models of retinal degeneration might be more suitable for studying retinal vasculature.

Animal and human studies may shed some light on the potential mechanism of peripheral retinal vascular loss in RP. Loss of retinal capillaries has been shown to correlate with the loss of photoreceptors in rodent models of RP ^19,20^. It has been postulated that loss of photoreceptors could increase oxygenation of the inner retina by a reduction in oxygen consumption in the outer retina as well as by an increased supply from the choroid ^7^. However, intraretinal oxygen tension measurements in a rat model of RP showed increased oxygen tension in the outer retina with the progression of photoreceptor degeneration but no major change in the oxygen tension of the inner retina ^21^. On the other hand, relative inner retinal hypoxia has been reported to be lower in a mouse model of RP compared to the wild-type, and this has been associated with decreased expression of hypoxia-regulated genes i.e. HIF-1α and vascular endothelial growth factor (VEGF) ^22^. Additionally, ambient hypoxia in a mouse model of RP has been shown to reverse the deep capillary plexus loss in this model ^23^. Human studies have demonstrated higher oxygen saturation in the retinal veins of RP patients compared to healthy controls suggesting decreased uptake of oxygen in the retina of these patients ^24,25^. Moreover, the concentration of VEGF-A in aqueous humor has been reported to be markedly lower in patients with RP than in control subjects ^26^.

Whether certain genotypes of RP are more susceptible to peripheral retinal vascular loss remains to be determined as our study was not powered to address this issue. However, based on the aforementioned animal studies, one could speculate that any such correlation would likely depend on the severity of photoreceptor degeneration in a given genotype.

Our findings in this study could have significant clinical implications. First, in addition to the classic findings of RP such as bone spicule pigmentation, vascular attenuation and waxy pallor of the optic disc, shortening of peripheral retinal vessels on UWF images may aid in clinical diagnosis of RP. Symmetrical loss of peripheral retinal vessels may even be a better clinical sign than vascular attenuation and disc pallor, which are subjective and may be difficult to appreciate. Of note, 5% of RP patients had 25 or more vessel crossings at Z2/3, indicating that a normal-appearing peripheral retinal vasculature cannot be used to rule out RP. Further studies are needed to evaluate the peripheral retinal vessels in other conditions, particularly non-RP inherited retinal diseases. Second, it appears that at least in some RP patients, a significant portion of the peripheral retina is solely nourished by the choroid, without any contribution from the retinal vessels. This is an important factor to keep in mind when creating subretinal blebs during stem cell transplantation or gene therapy; a creation or extension of the subretinal bleb beyond retinal vessels may leave that part of the retina without blood supply and risk ischemia or necrosis. Third, provided future studies demonstrate a correlation between the disease severity and the loss of peripheral retinal vessels, the length of retinal vessels could be used to monitor the progression of RP. Fourth, although previous studies point to photoreceptor loss as a cause for the loss of retinal vessels, it is plausible that decreased retinal blood supply may exacerbate photoreceptor degeneration. Therefore, potential measures that could prevent retinal vascular degeneration without inducing local hyperoxia, may slow down the progression of the disease.

One limitation of this study is its retrospective nature, which may introduce selection bias. Additionally, while UWF imaging provides extensive coverage of the retina, it still does not cover the entire retinal periphery. Future prospective studies with imaging of the entire retinal periphery could allow mapping of the avascular retinal periphery to explore the relationship between vascular loss and functional outcomes such as visual field changes. An additional limitation of this study is the limited visibility of small vessels on color fundus photos, as we demonstrated there were more visibility of vessels on FA. However, even FA images showed a significantly low vascularity index in RP, and given the non-invasive nature of fundus photos they could still be useful for studies of this kind.

In conclusion, our study provides compelling evidence of significant peripheral retinal vascular loss in RP. This finding may aid in clinical diagnosis of the disease. It may also have significant therapeutic implications in retinal gene therapy and stem cell transplantation. It also suggests potential targets for future therapeutic strategies aimed at preserving retinal vasculature and slowing disease progression.

## Supporting information

Supplementary ables

## Data Availability

All data produced in the present study are available upon reasonable request to the authors

## Acronyms

RP: retinitis pigmentosa
UWF: ultra-wide field
FA: fluorescein angiogram
IRR: incidence rate ratio
GEE: generalized estimating equation
CI: confidence interval
VA: visual acuity
LogMAR: Logarithmic Minimum Angle of Resolution
QIC: Quasi-Information Criterion

## Notes

Financial Support: This work was supported by grant UL1TR001855 from the National Center for Advancing Translational Science (NCATS) of the U.S. National Institutes of Health. The content is solely the responsibility of the authors and does not necessarily represent the official views of the National Institutes of Health. The Department of Ophthalmology received an unrestricted grant from Research to Prevent Blindness (New York, NY). The sponsor or funding organization had no role in the design or conduct of this research.

### Competing Interest Statement

The authors have declared no competing interest.

### Funding Statement

This work was supported by grant UL1TR001855 from the National Center for Advancing Translational Science (NCATS) of the U.S. National Institutes of Health. The content is solely the responsibility of the authors and does not necessarily represent the official views of the National Institutes of Health. The Department of Ophthalmology received an unrestricted grant from Research to Prevent Blindness (New York, NY). The sponsor or funding organization had no role in the design or conduct of this research.

### Author Declarations

The study was approved by the Institutional Review Board of the University of Southern California.

