## Supplementary Figure 1 for "Loss of Peripheral Retinal Vessels in Retinitis Pigmentosa"

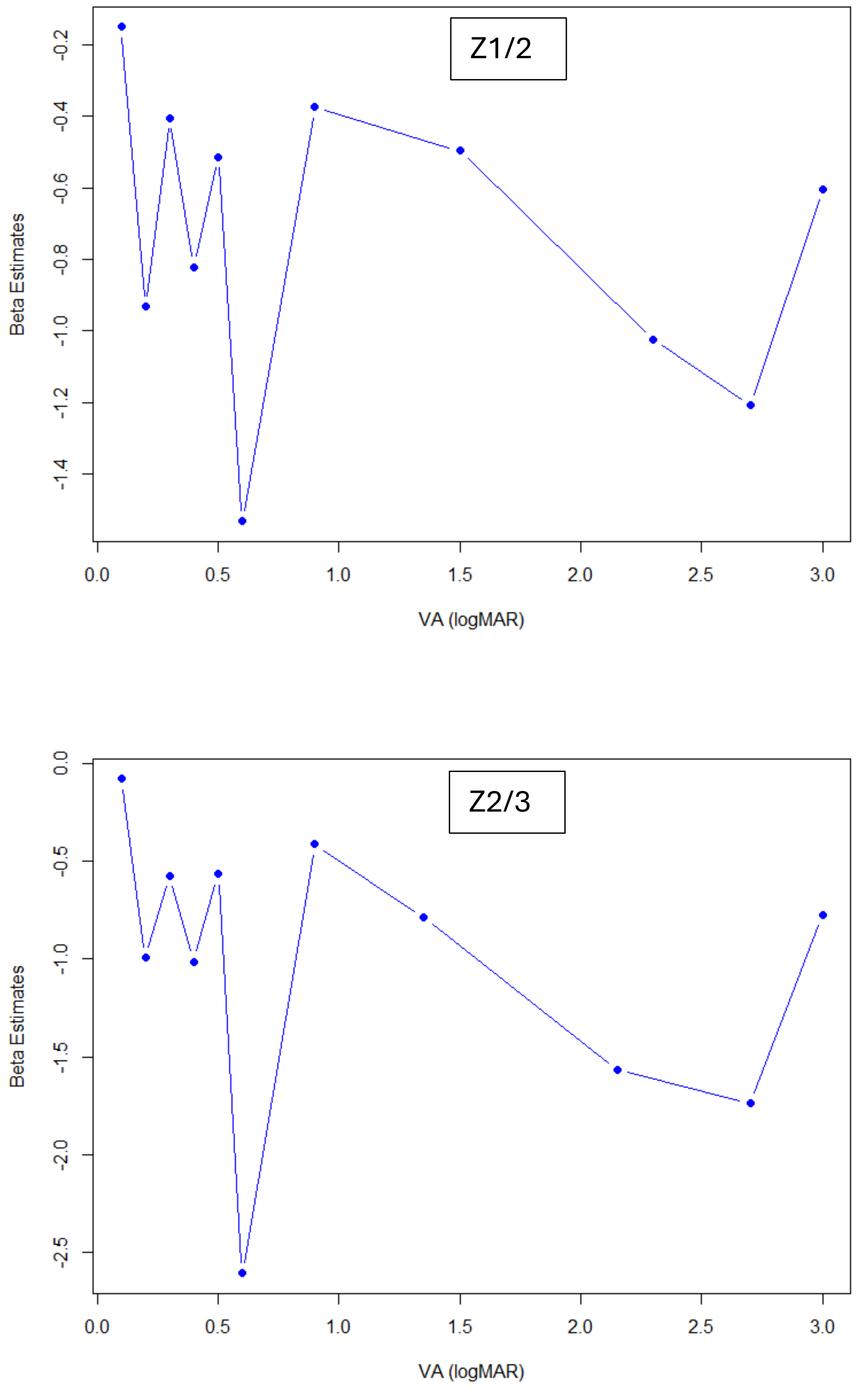


**Supplementary Figure 1**: Linearity assessment of grouped logMAR visual acuity in relation to total vessel counts in RP patients at Z1/2 (top) and Z2/3 (bottom) with beta coefficients from GEE model adjusting for age quartiles and sex. Both plots suggest non-linearity.
