## Supplementary ables for "Loss of Peripheral Retinal Vessels in Retinitis Pigmentosa"

**Supplementary Table 1**. Comparison of adjusted incidence rate ratio (IRR) for different quadrants at Z1/2 and Z2/3 in the control and retinitis pigmentosa (RP) groups. CI: confidence interval, REF: reference, IN: inferonasal, IT: inferotemporal, SN: superonasal, ST: superotemporal

| | | Z1/2 | | | Z2/3 | | |
|---|---|---|---|---|---|---|---|
| Variable | | IRR | 95% CI | P Value | IRR | 95% CI | P Value |
| Factor * Quadrant | | | | <0.001 | | | <0.001 |
| RP | IN | REF | | | REF | | |
| | IT | 0.78 | 0.65, 0.94 | 0.05 | 0.54 | 0.39, 0.75 | 0.001 |
| | SN | 1.27 | 1.16. 1.39 | <0.001 | 1.36 | 1.21, 1.52 | <0.001 |
| | ST | 1.12 | 0.97, 1.30 | 0.39 | 0.78 | 0.59, 1.04 | 0.33 |
| Control | IN | REF | | | REF | | |
| | IT | 1.64 | 1.56, 1.72 | <0.001 | 1.48 | 1.41, 1.56 | <0.001 |
| | SN | 1.13 | 1.08, 1.18 | <0.001 | 1.09 | 1.04, 1.14 | 0.002 |
| | ST | 1.69 | 1.61, 1.77 | <0.001 | 1.34 | 1.27, 1.41 | <0.001 |
| Age | | | | | | | |
| 14-33 | | REF | | | REF | | |
| 34-47 | | 0.94 | 0.84, 1.06 | 0.31 | 0.92 | 0.80, 1.07 | 0.27 |
| 48-63 | | 0.83 | 0.75, 0.93 | 0.001 | 0.81 | 0.72, 0.92 | 0.002 |
| 64-86 | | 0.74 | 0.66, 0.83 | <0.001 | 0.64 | 0.57, 0.73 | <0.001 |
| Sex | | | | | | | |
| Female | | REF | | | REF | | |
| Male | | 1.09 | 1.00, 1.19 | 0.06 | 1.08 | 0.97, 1.20 | 0.17 |

**Supplementary Table 2**. Comparison of adjusted incidence rate ratio (IRR) for horizontal hemispheres at Z1/2 and Z2/3 in the control and retinitis pigmentosa (RP) groups. CI: confidence interval, REF: reference

| | | Z1/2 | | | Z2/3 | | |
|---|---|---|---|---|---|---|---|
| Variable | | IRR | 95% CI | P Value | IRR | 95% CI | P Value |
| Factor * Horizontal Hemispheres | | | | <0.001 | | | <0.001 |
| RP | Inferior | REF | | | REF | | |
| | Superior | 1.34 | 1.23, 1.46 | <0.001 | 1.38 | 1.22, 1.57 | <0.001 |
| Control | Inferior | REF | | | REF | | |
| | Superior | 1.07 | 1.03, 1.11 | <0.001 | 0.98 | 0.94, 1.02 | 0.27 |
| Age | | | | | | | |
| 14-33 | | REF | | | REF | | |
| 34-47 | | 0.97 | 0.82, 1.15 | 0.72 | 1.13 | 0.89, 1.43 | 0.32 |
| 48-63 | | 0.81 | 0.69, 0.95 | 0.01 | 0.89 | 0.71, 1.11 | 0.32 |
| 64-86 | | 0.68 | 0.58, 0.79 | <0.001 | 0.63 | 0.52, 0.76 | <0.001 |
| Sex | | | | | | | |
| Female | | REF | | | REF | | |
| Male | | 1.07 | 0.95, 1.22 | 0.27 | 1.03 | 0.86, 1.23 | 0.74 |

**Supplementary Table 3**. Comparison of adjusted incidence rate ratio (IRR) for vertical hemispheres at Z1/2 and Z2/3 in the control and retinitis pigmentosa (RP) groups. CI: confidence interval, REF: reference

| | | Z1/2 | | | Z2/3 | | |
|---|---|---|---|---|---|---|---|
| Variable | | IRR | 95% CI | P Value | IRR | 95% CI | P Value |
| Factor * Vertical Hemispheres | | | | <0.001 | | | <0.001 |
| RP | Nasal | REF | | | REF | | |
| | Temporal | 0.84 | 0.73, 0.96 | 0.01 | 0.54 | 0.41, 0.72 | <0.001 |
| Control | Nasal | REF | | | REF | | |
| | Temporal | 1.56 | 1.51, 1.62 | <0.001 | 1.35 | 1.30, 1.40 | <0.001 |
| Age | | | | | | | |
| 14-33 | | REF | | | REF | | |
| 34-47 | | 0.97 | 0.81, 1.16 | 0.76 | 1.20 | 0.92, 1.57 | 0.18 |
| 48-63 | | 0.80 | 0.68, 0.95 | 0.01 | 0.93 | 0.72, 1.19 | 0.54 |
| 64-86 | | 0.68 | 0.58, 0.80 | <0.001 | 0.64 | 0.52, 0.79 | <0.001 |
| Sex | | | | | | | |
| Female | | REF | | | REF | | |
| Male | | 1.06 | 0.93, 1.21 | 0.35 | 1.02 | 0.83, 1.24 | 0.88 |

**Supplementary Table 4**. Comparison of adjusted incidence rate ratio (IRR) for the right and left eyes in the retinitis pigmentosa (RP) group with bilateral eyes at Z1/2 and Z2/3. CI: confidence interval, REF: reference

| | Z1/2 | | | Z2/3 | | |
|---|---|---|---|---|---|---|
| Variable | IRR | 95% CI | P Value | IRR | 95% CI | P Value |
| Eye | | | | | | |
| Right | REF | | | REF | | |
| Left | 0.95 | 0.85, 1.06 | 0.39 | 1.04 | 0.90, 1.20 | 0.56 |
| Age | | | | | | |
| 14-33 | REF | | | REF | | |
| 34-47 | 0.97 | 0.59, 1.59 | 0.91 | 1.21 | 0.60, 2.44 | 0.59 |
| 48-63 | 0.77 | 0.49, 1.23 | 0.28 | 0.94 | 0.48, 1.85 | 0.87 |
| 64-86 | 0.46 | 0.29, 0.74 | 0.001 | 0.38 | 0.20, 0.72 | 0.003 |
| Sex | | | | | | |
| Female | REF | | | REF | | |
| Male | 0.95 | 0.66, 1.37 | 0.80 | 0.78 | 0.46, 1.34 | 0.37 |

**Supplementary Table 5**. Comparison of adjusted incidence rate ratio (IRR) for the right and left eyes in the control group with bilateral eyes at Z1/2 and Z2/3. CI: confidence interval, REF: reference

| | Z1/2 | | | Z2/3 | | |
|---|---|---|---|---|---|---|
| Variable | IRR | 95% CI | P Value | IRR | 95% CI | P Value |
| Eye | | | | | | |
| Right | REF | | | REF | | |
| Left | 0.96 | 0.92, 1.00 | 0.03 | 1.00 | 0.95, 1.05 | 1.00 |
| Age | | | | | | |
| 14-33 | REF | | | REF | | |
| 34-47 | 0.95 | 0.85, 1.06 | 0.33 | 0.92 | 0.83, 1.01 | 0.08 |
| 48-63 | 0.92 | 0.83, 1.01 | 0.09 | 0.94 | 0.85, 1.04 | 0.21 |
| 64-86 | 0.91 | 0.82, 1.01 | 0.08 | 0.82 | 0.75, 0.89 | <0.001 |
| Sex | | | | | | |
| Female | REF | | | REF | | |
| Male | 1.07 | 0.98, 1.17 | 0.12 | 1.08 | 1.00, 1.17 | 0.05 |

**Supplementary Table 6**. Sensitivity analysis of adjusted incidence rate ratio (IRR) of logMAR visual acuity in retinitis pigmentosa patients. This analysis is similar to table 6, except that the cut-off logMAR visual acuity is 0.7 instead of 0.47. CI: confidence interval, REF: reference

| | Z1/2 | | | Z2/3 | | |
|---|---|---|---|---|---|---|
| Variable | IRR | 95% CI | P Value | IRR | 95% CI | P Value |
| Visual Acuity (logMAR) | | | | | | |
| 0.0-0.69 | REF | | | REF | | |
| 0.70-0.99 | 1.41 | 0.73, 2.73 | 0.31 | 1.51 | 0.67, 3.43 | 0.32 |
| 1.0-3.0 | 0.63 | 0.47, 0.83 | 0.001 | 0.42 | 0.27, 0.66 | <0.001 |
| Age | | | | | | |
| 14-33 | REF | | | REF | | |
| 34-47 | 1.08 | 0.80, 1.47 | 0.61 | 1.67 | 1.11, 2.53 | 0.01 |
| 48-63 | 0.78 | 0.57, 1.06 | 0.11 | 1.04 | 0.68, 1.60 | 0.86 |
| 64-86 | 0.64 | 0.44, 0.91 | 0.01 | 0.68 | 0.41, 1.12 | 0.13 |
| Sex | | | | | | |
| Female | REF | | | REF | | |
| Male | 1.01 | 0.80, 1.29 | 0.91 | 0.96 | 0.68, 1.35 | 0.81 |
